# Associations of novel blood-derived markers of inflammation with blood pressure, arterial stiffness and heart rate in young adults

**DOI:** 10.1101/2025.06.17.25329810

**Authors:** Richard J Woodman, Angelo Zinellu, Trevor A Mori, Lawrie J Beilin, Arduino A Mangoni

## Abstract

**Background:** Although C-reactive protein (CRP) is often used to assess inflammation and can predict higher BP and arterial stiffness, novel measures of inflammation derived from platelets, white blood cell (WBC) counts, and high-density lipoprotein cholesterol (HDL-C) may also indicate possible hypertension and arterial stiffening.

**Methods:** We assessed the association for CRP and novel inflammatory markers with clinic BP, arterial stiffness, and heart rate (HR) in Gen2 Raine Study participants in Western Australia at age 17 and 22 years. Arterial stiffness was assessed using pulse-wave velocity (PWV), the augmentation index (AIx) and pulse pressure (PP). Inflammatory markers included high-sensitivity (hs)-CRP, hsCRP-to-albumin ratio, lymphocyte, monocyte, neutrophil, and platelet counts, neutrophil-to-lymphocyte ratio, platelet-to-lymphocyte ratio, mean platelet volume, the neutrophil percentage-to-albumin ratio, the monocyte/HDL-C ratio, the neutrophil/HDL-C ratio, the Prognostic Nutritional Index and the Systemic Inflammation Index (SII). Analysis adjusted for age, gender, waist-to-hip ratio, family history of hypertension, diet, physical activity, smoking and alcohol consumption.

**Results:** Males (N=363) had higher systolic BP (Δ=9.1mmHg; 95% CI=7.9-10.3, p<0.001), lower diastolic BP (Δ=−0.9mmHg; 95% CI=−1.8, −0.1, p<0.001) and higher PWV (Δ=0.37m/sec; 95% CI=0.26 to 0.48, p<0.001) than females (N=330). Lymphocyte count, the monocyte/HDL-C ratio and the neutrophil/HDL-C ratio were each significantly and positively associated with SBP, PP and the AIx. Platelet count was significantly and positively associated with SBP, DBP and PP. Nine of the 14 inflammatory indices were associated with HR, and associations for SBP, DBP, AIx and PWV were mostly stronger for males than for females. hsCRP and the SII were not associated with any of SBP, DBP, PP, AIx or PWV.

**Conclusions:** Lymphocyte count, the monocyte/HDL-C ratio, the neutrophil/HDL-C ratio, and platelet count may represent better markers of inflammation related to higher BP in young adults than CRP and the SII. The positive associations with arterial stiffness and HR, and the stronger associations for males also provides insight into potential mechanisms.

## 1. Introduction

Systemic inflammation is closely associated with vascular alterations, including endothelial dysfunction and increased arterial stiffness (1) and traditional markers of inflammation, e.g., C-reactive protein (CRP), have been associated with hypertension in large-scale studies (2, 3). In older adults, even low-grade inflammation is associated with cardiometabolic multimorbidity (4) and in healthy younger adults, inflammatory markers including cytokines were associated with increased blood pressure (BP) over 4.5 years (5), demonstrating that excess inflammation can exert significant negative effects on BP at an early age. Identification of easily attainable inflammatory markers for predicting BP elevations in young age would facilitate more targeted monitoring and interventions to prevent future hypertension and cardiovascular disease. For example, a modified intake of dietary fatty acids, salt, and ultra-processed foods which are each associated with increased inflammation and BP (6-8), might reduce the incidence of young adult hypertension.

Although inflammation predicts some forms of hypertension and the development of hypertension, there is no consensus on which marker of inflammation is best for this purpose (9). Most studies addressing the relationship between inflammation and BP have measured CRP as a non-specific marker of inflammation (2, 10) but many additional inflammatory markers, including neutrophil, lymphocyte, monocyte, and platelet counts, can be derived from routine hematological blood tests. Others include the neutrophil-lymphocyte ratio (11), lymphocyte-monocyte ratio (12), platelet-lymphocyte ratio (12), mean platelet volume (13) and the systemic inflammation index (SII) (14). Albumin and HDL-cholesterol (HDL-C) have also been used to normalize some of these counts to provide additional potential markers including the monocyte-to-HDL-C ratio (15), the neutrophil-to-HDL-C ratio (16), the CRP-to-albumin ratio (17) and the neutrophil percentage-to-albumin (18). WBC count measures may also be more specific markers of hypertension related inflammation since T-lymphocytes appear essential for the development of hypertension and the central nervous system contributes to hypertension via mechanisms including peripheral T-lymphocyte activation and vascular inflammation (1, 19). T-lymphocytes and vascular inflammation also contribute to stress-dependent hypertension (1). Elevated monocyte-to-HDL and neutrophil-to-HDL ratios suggest a higher inflammatory state relative to protective HDL-C and are associated with increased risk of cardiovascular events (16, 20). The CRP-to-albumin (17) and neutrophil-percent-to-albumin (18) ratios combine an inflammatory marker with a measure of nutritional status and have been studied in cardiovascular and kidney disease. However, there is relatively little information regarding whether these indices might better predict BP elevations than CRP in young adults.

Our primary aim was to investigate associations between CRP, CRP-to-albumin, twelve novel markers of inflammation (Neutrophil count, Lymphocyte count, Monocyte count, Platelet count, Neutrophil-to-lymphocyte ratio, Platelet-to-lymphocyte ratio, Monocyte-to-platelet volume, SII, monocyte-to-HDL ration, Neutrophil-to-HDL ratio, neutrophil percent-to-albumin and PNI), and systolic BP (SBP) and diastolic BP (DBP). Our secondary aims were to investigate whether these blood count derived inflammatory indices and CRP are associated with direct and indirect measures of arterial stiffness (pulse pressure, (PP), augmentation index (AIx), and pulse-wave velocity, (PWV) and heart rate (HR) to assess potential mechanisms. We obtained repeat cross-sectional measures of blood counts and clinic BP at ages 17 and 22 years, and AIx and PWV obtained at age 17, in a population of healthy males and females from the Raine Study.

## 2. Materials and Methods

### 2.1 Participants

The Raine Study (www.rainestudy.org.au) represents a large cohort of Western Australian offspring studied from 18 weeks’ gestation to ascertain the relative contributions of familial risk factors, fetal growth, placental development, and environmental insults to outcome in infancy and to the precursors of adult morbidity. A total of 2900 women (Gen1) were enrolled into the study from 1989-1991. There were 2868 live births (Gen2), that have been prospectively followed up at regular intervals from birth, with demographic, lifestyle, clinical and biochemical information collected through questionnaires and clinical assessments. (21). The Human Ethics Committees at King Edward Memorial Hospital, Princess Margaret Hospital for Children, and the University of Western Australia in Perth approved all recalls of the cohort. This study included N=692 Gen2 participants with clinic BP data and inflammatory markers available at both age 17 and 22. Their selection is described in further detail in a CONSORT flow chart (Figure S1).

### 2.2 Demographic and clinical data

Demographic data including smoking status (yes/no), level-of-physical-activity (low/moderate/high), use of oral/hormonal contraceptives (at age 17 and 22) and alcohol consumption by age 17 (none/just-a-few-sips/less-than-10-sips/more-than-10-sips) were recorded in medical questionnaires. An international physical activity questionnaire (IPAQ) recorded metabolic equivalents (METs) at age 17 and age 22. Clinical assessments included waist-to-hip ratio (WHR). Information on a family history of high BP in either the mother or the father was recorded at age 8. Figure S1 provides details on the number of individuals assessed for each measurement at 17- and 22-years, and the extent of missing information.

### 2.3 Clinic BP and heart rate

Resting clinic BP and HR were measured after an overnight fast with an appropriate cuff size for arm circumference, using a semi-automated oscillometric monitor (DINAMAP ProCare 100 vital signs monitor; GE Healthcare, USA). Recordings were obtained from the right arm after the participant had been seated for ≥ 5 mins. Six measurements were taken 2-minutes apart and the first recording was removed from the analysis (22) before averaging the remaining 5 readings for analysis.

### 2.4 Arterial stiffness

Measures of arterial stiffness included PWV, the AIx, and PP, calculated as mean systolic BP minus mean diastolic BP. The AIx was measured using pulse wave analysis (PWA) (Sphygmocor, software version 1.2, AtCor Medical Pty Ltd, Sydney, Australia). PWA data was collected from a supported radial artery with the wrist facing upward and data captured after a consistent waveform was maintained for 10 seconds. The test was repeated until at least 2 captures were recorded with a quality index of >80. AIx was defined as the difference in the second and first systolic pressure peaks as a percentage of pulse pressure and was corrected to a heart rate of 75 bpm using AIx@ HR75=[−0.48×(75−HR)]+AIx (23). For the PWV, 3 ECG leads were attached to the left leg, right arm, and left arm. Tonometers were applied to the carotid artery and distal dorsalis pedis and the distance (mm) was taken between the manubrium sternum and the 2 sampling sites. PWV was calculated by dividing the distance between the tonometers by the transit time of the arterial pulse wave (24). The day-to-day coefficient of variation for PWV was 5%.

### 2.5 Inflammatory markers

The 14 inflammatory markers studied included high-sensitivity CRP, CRP/Albumin, Lymphocytes, Monocytes, Neutrophils, Platelets, Neutrophil-to-Lymphocyte Ratio, Platelet-to-Lymphocyte Ratio, Monocyte-to-Platelet Volume, Neutrophil-percent/Albumin ratio, Monocyte/HDL-C ratio, Neutrophil/HDL-C ratio, the Prognostic Nutritional Index (PNI) and the Systemic Inflammation Index (SII) calculated as [lymphocyte count/platelet count] x neutrophil count (25). PNI was assessed using [5 × lymphocyte count (10^9^/L)] + serum albumin (g/L). Serum hsCRP, stored at −80°C within 2h of collection was analysed using an immunoturbidimetric method on an Architect c16000 Analyzer (Abbott core Laboratory, Illinois, United States of America). A single aliquot of whole blood was processed for full blood count, including circulating monocytes, neutrophils, lymphocytes and platelets. HDL-C was determined on a heparin–manganese supernatant (26). High-sensitivity CRP (hsCRP), HDL-C and blood counts were analysed by PathWest Laboratory (Royal Perth Hospital) (27).

### 2.6 Dietary intake and alcohol consumption

Information on dietary intake, which was used in regression adjustment, was estimated using the 74-item semiquantitative Dietary Questionnaire for Epidemiological Studies (DQESV2) FFQ, developed by the Cancer Council of Victoria, Australia (28). Standardized scores for Healthy and Western diets at age 17 were calculated using factor analysis of major food group intakes obtained from a semi-quantitative dietary recall questionnaire designed by Australia’s Commonwealth Scientific and Industrial Research Organization (CSIRO) (29, 30). These two extracted factors explained 13% (Western dietary pattern) and 8.5% (Healthy dietary pattern) of the total variance in food intake (31). Additional information recorded from the CSIRO dietary food recall records included sodium and fibre intake, frequency of fruit and vegetable consumption, and the standard number of alcoholic drinks consumed per week captured from a medical questionnaire at age 20. This was recorded both as grams of alcohol/day for a typical week and using 3 categories (<1 standard drink/day, 1-3 standard drinks/day and more than 3 standard drinks/day).

### 2.7 Statistical analysis

The study population was described using mean±SD for normally distributed variables, median (inter-quartile range) for non-normally distributed variables, and frequencies (percentages) for categorical variables. Differences between males and females were assessed using an independent t-test or Mann-Whitney U-test for continuous variables, and a chi-squared test for frequency data. Associations between inflammatory markers and clinic systolic BP, diastolic BP, PP and HR were assessed using a linear mixed effects model with the subject ID included as a random intercept, and an unstructured covariance matrix for the residuals. The random effect accounted for the repeated measures on the same individual at age 17 and age 22. The associations between inflammatory markers and AIx and PWV (measured at age 17 only) were assessed using a generalized linear model with Gaussian distribution and identity link. The exposure of interest for each model was one of the fourteen individual inflammatory markers, included as a continuous variable without transformation. The dependent variables were mean clinic SBP, mean clinic DBP, mean clinic PP, mean clinic HR, AIx and PWV. Associations were determined with and without adjustment for potential confounders which included age, waist-to-hip ratio, family history of hypertension, healthy and western diet factor analysis scores, frequency of fruit and vegetable consumption, alcohol intake (g/day), sodium, and fibre intake, METs per week, and physical activity category (0/1/2). All regression model results are reported as the mean change (β) (95% confidence interval) for a one standard deviation increase in each inflammatory marker. Due to missing data for some of the covariates (Figure S1), multiple imputation with chained equations (MICE) was used to create 20 multiply imputed datasets which were used for all analyses. The MICE routine used predictive mean matching to impute categorical variables and linear regression to impute continuous covariates. Non-missing variables included in the imputation were systolic and diastolic BP measurements, participant ID and visit (17 versus 22 years). All analyses were performed using Stata (version 17.0, StataCorp, U.S.A.).

## 3. Results

### 3.1 Study population

From the original 2868 Gen2 participants there were 693 participants with complete data on inflammatory markers and clinic BP at age 17 and at age 22. The CONSORT flow diagram (Figure S1) describes the inclusion and exclusion of subjects from the original cohort.

Table 1 describes the demographic and lifestyle characteristics of the 693 participants. There were slightly more males (n=363, 52.4%) than females (n=330, 47.6%), and the overall mean (±SD) WHR across all participants was 0.81±0.07 at age 17 and 0.83±0.07 at age 22, with females having a lower WHR than males at age 17 and at age 22 (p<0.001). Participants were generally meeting fruit, vegetable, and alcohol recommendations at age 17 and 22 according to the Australian Guide to Healthy Eating (AGHE). Western diet factor scores and healthy diet factor scores at age 17 were comparable to the full Raine Study Gen2 cohort with means (±SD) of −0.05±0.82 and 0.05±0.89 respectively and close to zero. However, females had a slightly healthier profile than males for the Western diet factor score (−0.35±0.70 versus 0.25±0.83, p<0.001) but were similar for the healthy diet factor score (0.12±0.90 versus −0.03±0.89, p=0.076). Median alcohol consumption at age 20 was slightly lower amongst females than males (7.1 (0.0-17.1) g/day versus 11.4 (0.0-30.0) g/day, p<0.001). Most participants (N=547, 84.8%) were no-smokers at age 22. Males were higher in physical activity category, and in mean MET mins/week at both age 17 and age 22 than females. Table S1 describes the inflammatory markers, BP, HR, AIx and PWV of the participants. Most of the study participants were normotensive (SBP<130mmHg and DBP<80mmHg)(32) at age 17 (94.4%) and at age 22 (83.6%), with the overall mean (±SD) SBP at age 17 being 117.3±9.5 for males and 108.6±9.3 mmHg for females (p<0.001). At age 22 the corresponding mean SBP was 123.3±10.6 and 113.7±9.3 mmHg (p<0.001). Amongst the inflammatory markers, the monocyte count (p=0.009 and p<0.001) and the monocyte/HDL-C ratio (p<0.001 for each) was higher in males than in females at age 17 and age 22 whilst hsCRP was higher in females than in males at both ages (p<0.001 for each), as was platelet count (p<0.001 for each).

**Table 1:**
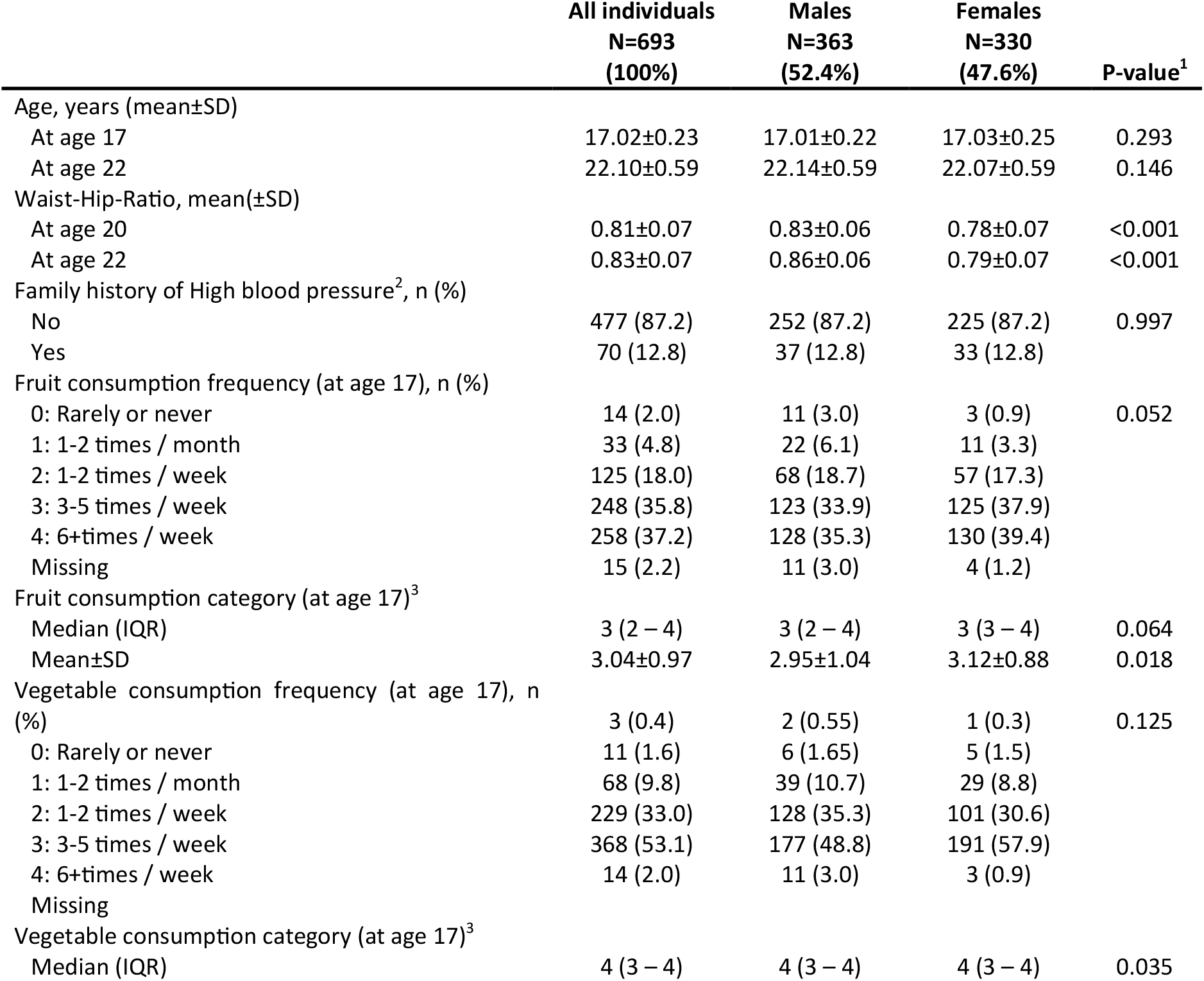
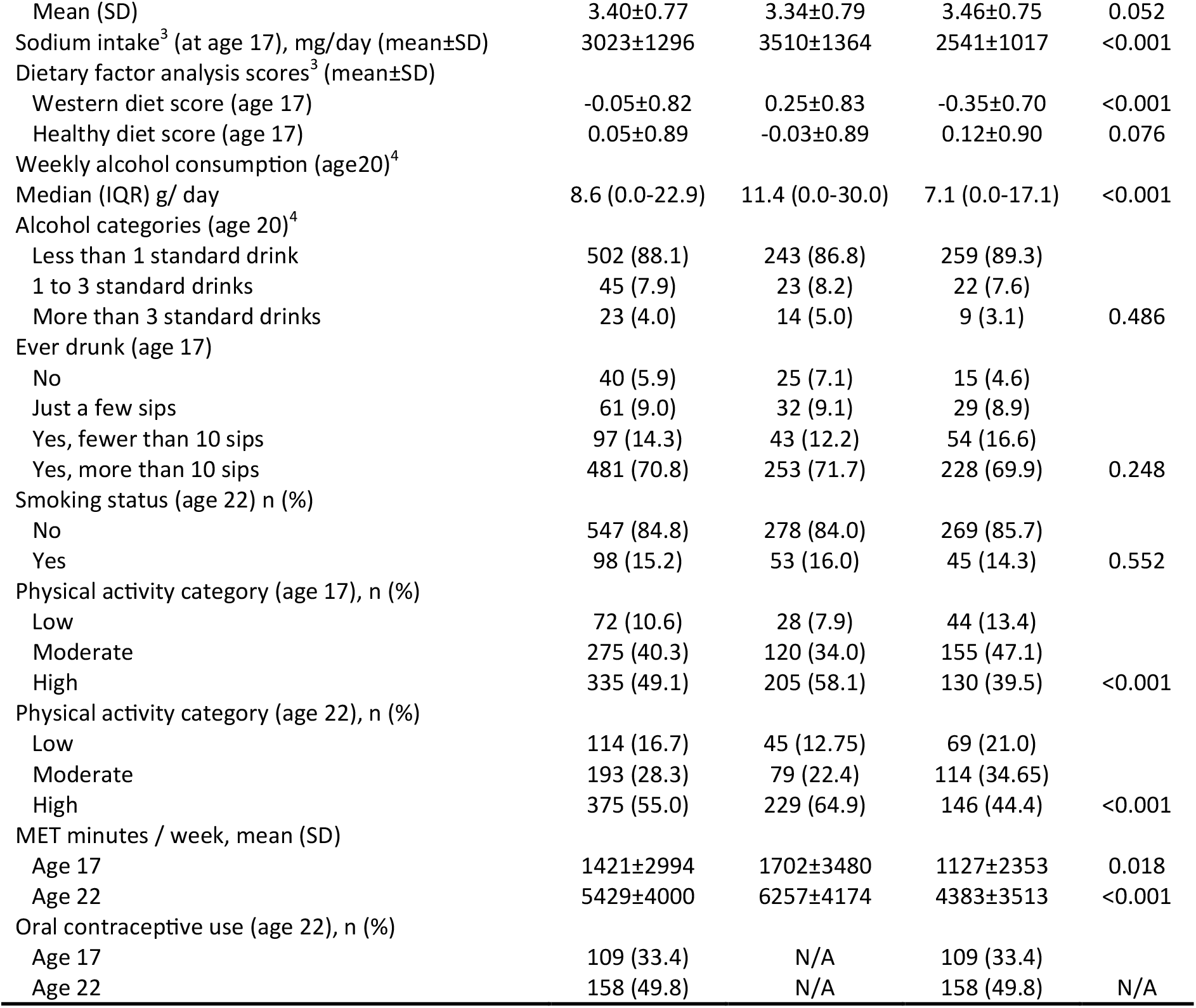
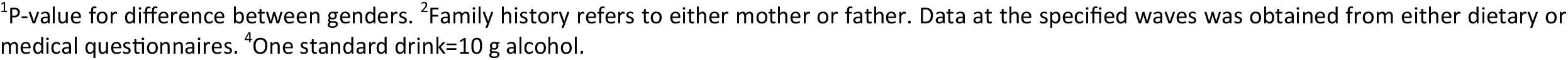
Demographics, diet, and lifestyle characteristics of Gen2 participants (N=693) at 17- and 22-years of age.

|  | **All individuals**<br>**N=693**<br>**(100%)** | **Males**<br>**N=363**<br>**(52.4%)** | **Females**<br>**N=330**<br>**(47.6%)** | **P-value<sup>1</sup>** |
| --- | --- | --- | --- | --- |
| Age, years (mean±SD) |  |  |  |  |
| At age 17 | 17.02±0.23 | 17.01±0.22 | 17.03±0.25 | 0.293 |
| At age 22 | 22.10±0.59 | 22.14±0.59 | 22.07±0.59 | 0.146 |
| Waist-Hip-Ratio, mean(±SD) |  |  |  |  |
| At age 20 | 0.81±0.07 | 0.83±0.06 | 0.78±0.07 | <0.001 |
| At age 22 | 0.83±0.07 | 0.86±0.06 | 0.79±0.07 | <0.001 |
| Family history of High blood pressure<sup>2</sup>, n (%) |  |  |  |  |
| No | 477 (87.2) | 252 (87.2) | 225 (87.2) | 0.997 |
| Yes | 70 (12.8) | 37 (12.8) | 33 (12.8) |  |
| Fruit consumption frequency (at age 17), n (%) |  |  |  |  |
| 0: Rarely or never | 14 (2.0) | 11 (3.0) | 3 (0.9) | 0.052 |
| 1: 1-2 times / month | 33 (4.8) | 22 (6.1) | 11 (3.3) |  |
| 2: 1-2 times / week | 125 (18.0) | 68 (18.7) | 57 (17.3) |  |
| 3: 3-5 times / week | 248 (35.8) | 123 (33.9) | 125 (37.9) |  |
| 4: 6+times / week | 258 (37.2) | 128 (35.3) | 130 (39.4) |  |
| Missing | 15 (2.2) | 11 (3.0) | 4 (1.2) |  |
| Fruit consumption category (at age 17)<sup>3</sup> |  |  |  |  |
| Median (IQR) | 3 (2 – 4) | 3 (2 – 4) | 3 (3 – 4) | 0.064 |
| Mean±SD | 3.04±0.97 | 2.95±1.04 | 3.12±0.88 | 0.018 |
| Vegetable consumption frequency (at age 17), n (%) |  |  |  |  |
| 0: Rarely or never | 3 (0.4) | 2 (0.55) | 1 (0.3) | 0.125 |
| 1: 1-2 times / month | 11 (1.6) | 6 (1.65) | 5 (1.5) |  |
| 2: 1-2 times / week | 68 (9.8) | 39 (10.7) | 29 (8.8) |  |
| 3: 3-5 times / week | 229 (33.0) | 128 (35.3) | 101 (30.6) |  |
| 4: 6+times / week | 368 (53.1) | 177 (48.8) | 191 (57.9) |  |
| Missing | 14 (2.0) | 11 (3.0) | 3 (0.9) |  |
| Vegetable consumption category (at age 17)<sup>3</sup> |  |  |  |  |
| Median (IQR) | 4 (3 – 4) | 4 (3 – 4) | 4 (3 – 4) | 0.035 |
| Mean (SD) | 3.40±0.77 | 3.34±0.79 | 3.46±0.75 | 0.052 |
| Sodium intake<sup>3</sup> (at age 17), mg/day (mean±SD) | 3023±1296 | 3510±1364 | 2541±1017 | <0.001 |
| Dietary factor analysis scores<sup>3</sup> (mean±SD) |  |  |  |  |
| Western diet score (age 17) | -0.05±0.82 | 0.25±0.83 | -0.35±0.70 | <0.001 |
| Healthy diet score (age 17) | 0.05±0.89 | -0.03±0.89 | 0.12±0.90 | 0.076 |
| Weekly alcohol consumption (age20)<sup>4</sup> |  |  |  |  |
| Median (IQR) g/ day | 8.6 (0.0-22.9) | 11.4 (0.0-30.0) | 7.1 (0.0-17.1) | <0.001 |
| Alcohol categories (age 20)<sup>4</sup> |  |  |  |  |
| Less than 1 standard drink | 502 (88.1) | 243 (86.8) | 259 (89.3) |  |
| 1 to 3 standard drinks | 45 (7.9) | 23 (8.2) | 22 (7.6) |  |
| More than 3 standard drinks | 23 (4.0) | 14 (5.0) | 9 (3.1) | 0.486 |
| Ever drunk (age 17) |  |  |  |  |
| No | 40 (5.9) | 25 (7.1) | 15 (4.6) |  |
| Just a few sips | 61 (9.0) | 32 (9.1) | 29 (8.9) |  |
| Yes, fewer than 10 sips | 97 (14.3) | 43 (12.2) | 54 (16.6) |  |
| Yes, more than 10 sips | 481 (70.8) | 253 (71.7) | 228 (69.9) | 0.248 |
| Smoking status (age 22) n (%) |  |  |  |  |
| No | 547 (84.8) | 278 (84.0) | 269 (85.7) |  |
| Yes | 98 (15.2) | 53 (16.0) | 45 (14.3) | 0.552 |
| Physical activity category (age 17), n (%) |  |  |  |  |
| Low | 72 (10.6) | 28 (7.9) | 44 (13.4) |  |
| Moderate | 275 (40.3) | 120 (34.0) | 155 (47.1) |  |
| High | 335 (49.1) | 205 (58.1) | 130 (39.5) | <0.001 |
| Physical activity category (age 22), n (%) |  |  |  |  |
| Low | 114 (16.7) | 45 (12.75) | 69 (21.0) |  |
| Moderate | 193 (28.3) | 79 (22.4) | 114 (34.65) |  |
| High | 375 (55.0) | 229 (64.9) | 146 (44.4) | <0.001 |
| MET minutes / week, mean (SD) |  |  |  |  |
| Age 17 | 1421±2994 | 1702±3480 | 1127±2353 | 0.018 |
| Age 22 | 5429±4000 | 6257±4174 | 4383±3513 | <0.001 |
| Oral contraceptive use (age 22), n (%) |  |  |  |  |
| Age 17 | 109 (33.4) | N/A | 109 (33.4) |  |
| Age 22 | 158 (49.8) | N/A | 158 (49.8) | N/A |

<sup>1</sup>P-value for difference between genders. <sup>2</sup>Family history refers to either mother or father. Data at the specified waves was obtained from either dietary or medical questionnaires. <sup>4</sup>One standard drink=10 g alcohol.

### 3.2 Inflammatory marker correlations

Table S2 describes the Spearman correlations between the 14 inflammatory markers and the covariates used for adjustment. Several variables including gender, WHR, Western diet score, MET mins/week and smoking status were consistently associated with the inflammatory markers including monocytes versus sex (ρ= −0.54, p<0.001) indicating a higher level of monocytes for males compared to females. Monocytes/HDL-C was associated with sex (ρ=−0.35) and WHR (ρ=0.33), and Neutrophils/HDL-C was associated with WHR (ρ=0.28) (p<0.05 for each). Figures S2 to S8 display scatter plots for the inflammatory markers versus SBP (Fig S2), age (Fig S3), sex (Fig S4), WHR (Fig S5), MET mins/week (Fig S6), western diet score (Fig S7), and smoking (Fig S8) respectively.

### 3.3 Regression analysis of inflammatory markers

Figures 1-3 and Figures S9-S11 show the strength of the unadjusted and fully adjusted associations between each inflammatory marker and SBP, DBP, AIx, HR, PP and PWV respectively. Tables S3 and S4 provide details of the fully adjusted associations (model 3), and the results of testing for effect modification between males and females. SBP was associated with Lymphocytes, Monocyte-to-HDL-C ratio, Neutrophil-to-HDL-C ratio, Monocyte count, Neutrophil count, Platelet count, and PNI. DBP was associated with Monocyte-to-platelet volume, Neutrophil-to-HDL-C ratio, Neutrophil count, Platelet count, and PNI. PP was associated with Lymphocyte count, Monocyte-to-HDL-C ratio, Neutrophil-to-HDL ratio, Monocyte count, and Platelet count. The AIx was associated with Lymphocytes, Monocyte-to-HDL ratio, and the Neutrophil-to-HDL ratio. PWV was associated with the Platelet/lymphocyte ratio.

**Figure 1:**
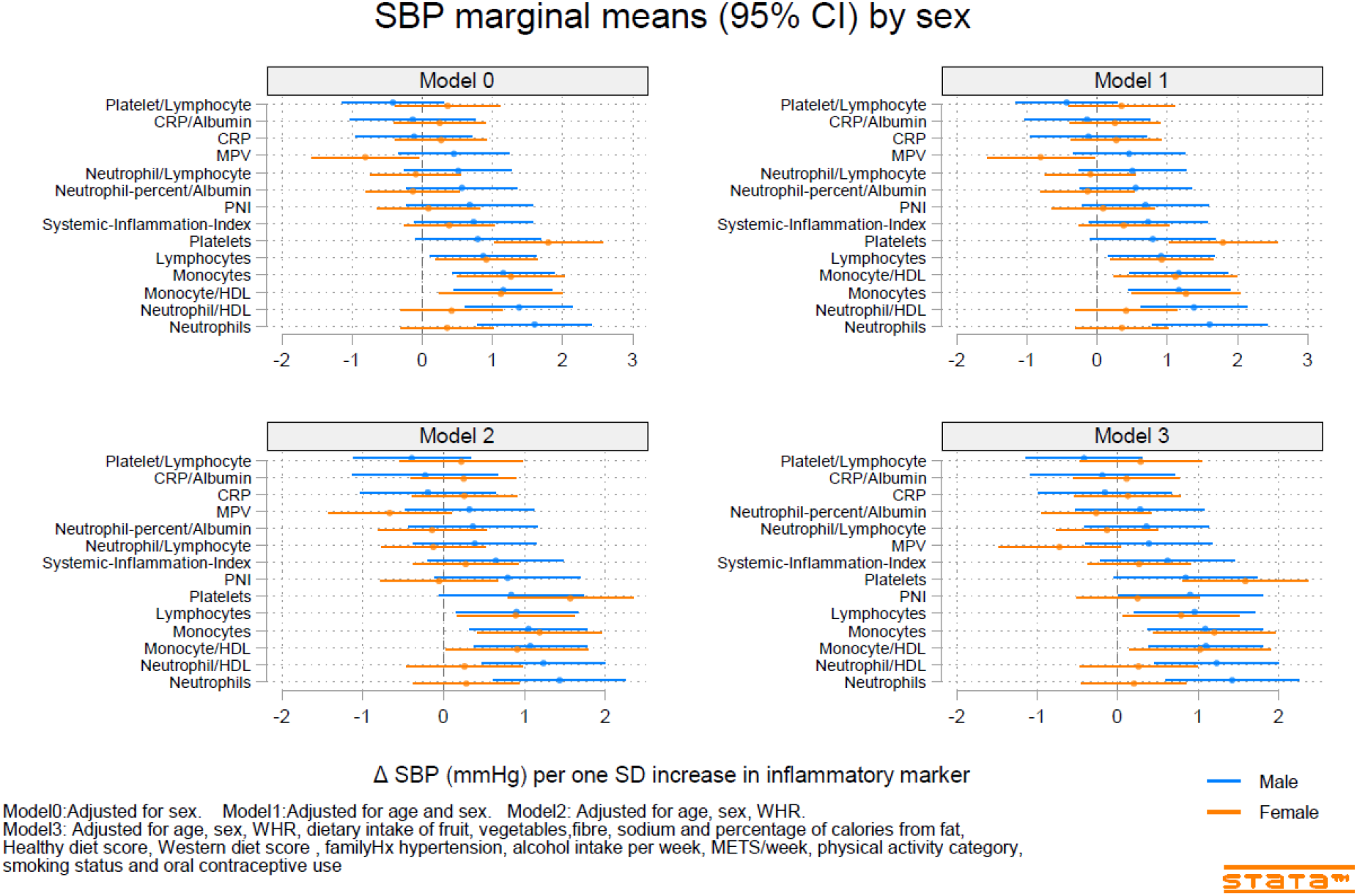
Predicted marginal mean effect of inflammatory markers on clinic systolic blood pressure for males (N=363) and females (N=330).

**Figure 2:**
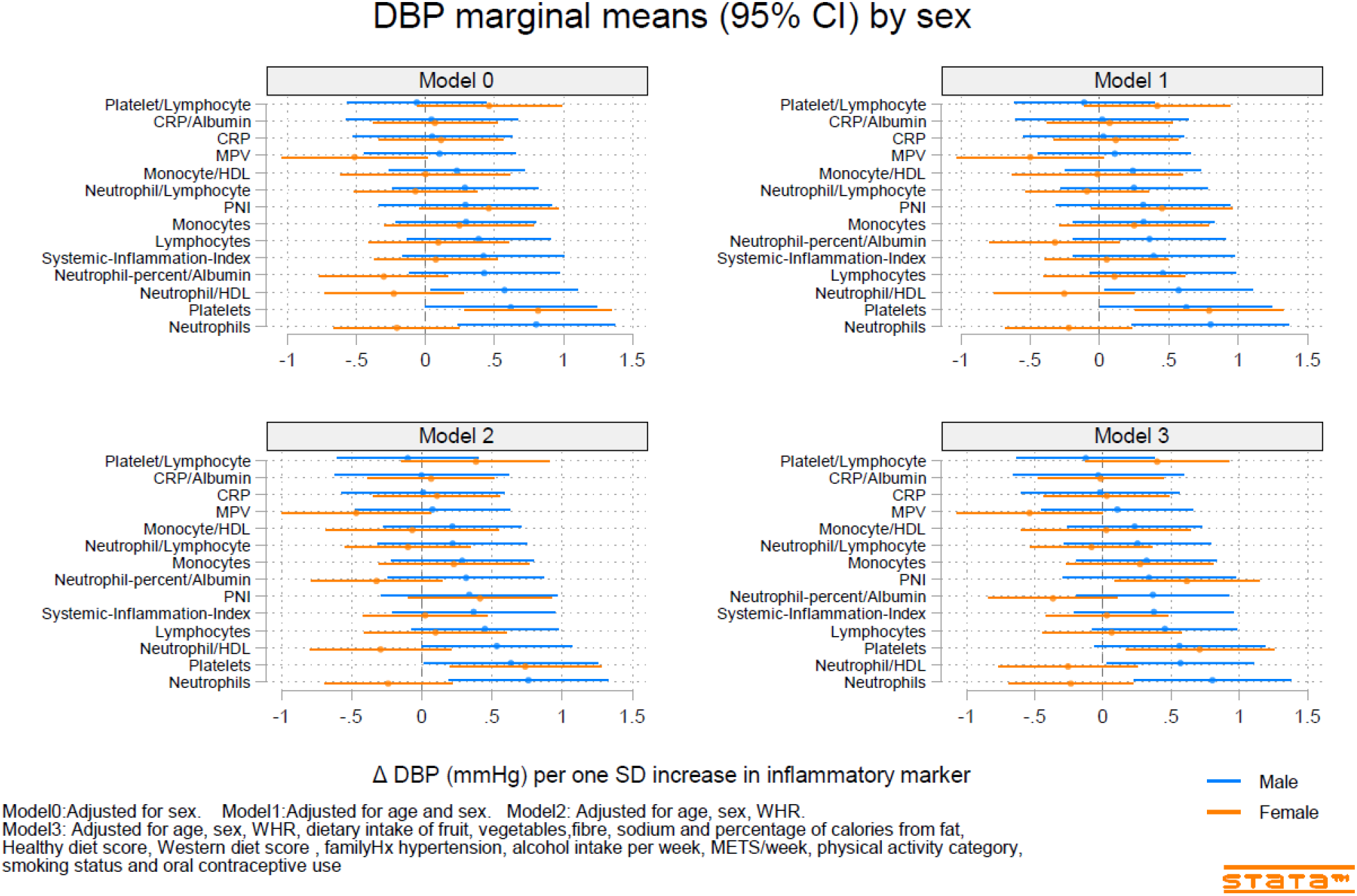
Predicted marginal mean effect of inflammatory markers on clinic diastolic blood pressure for males (N=363) and females (N=330).

**Figure 3:**
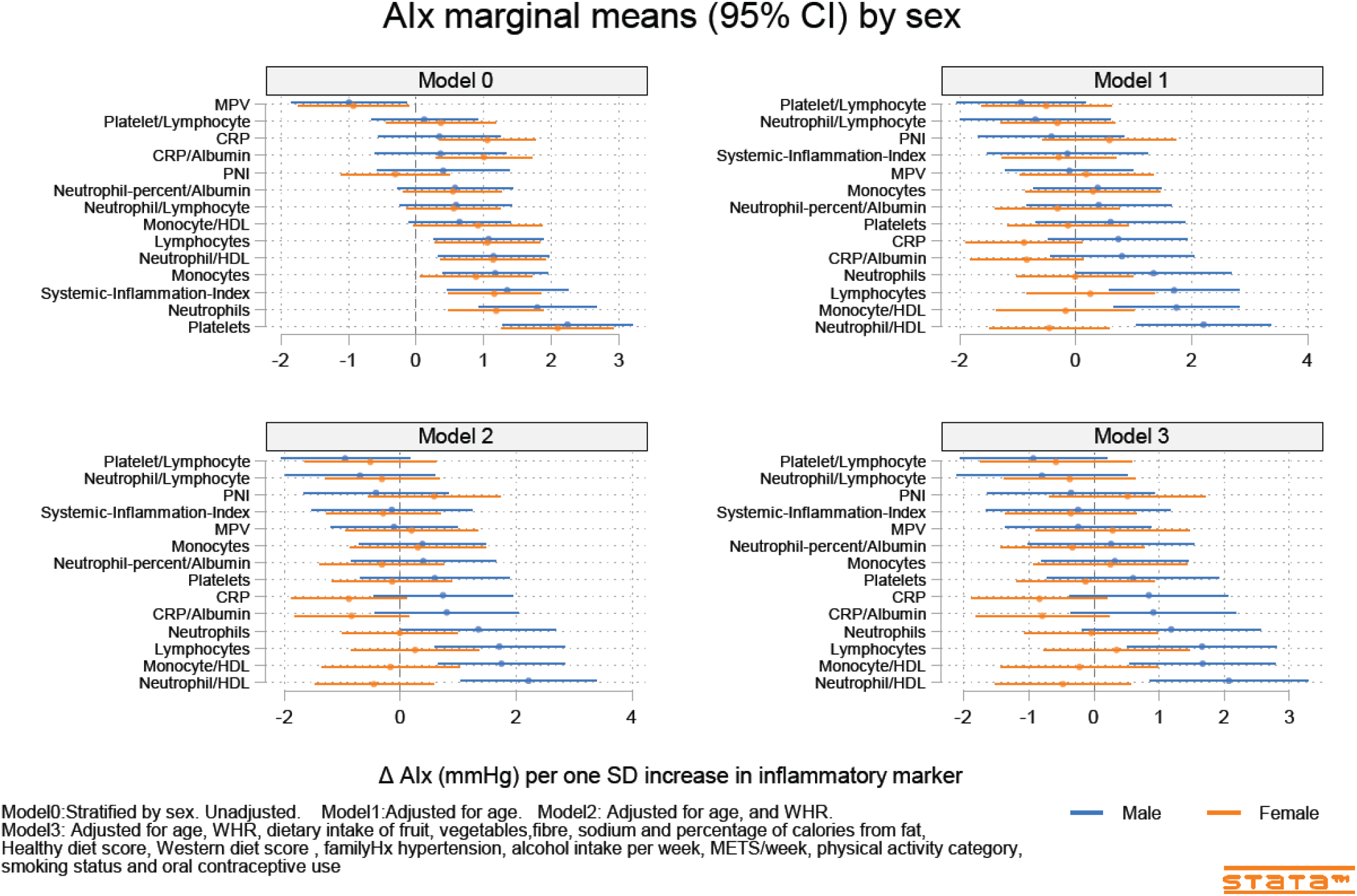
Predicted marginal mean effects of inflammatory markers on AIx for males (N=344) and females (N=301).

The most consistently associated inflammatory markers included Lymphocyte count, the Monocyte-to-HDL-C ratio and the Neutrophil-to-HDL-C ratio which were each significantly associated with SBP, PP and the AIx. Platelet count was significantly associated with SBP, DBP and PP. Amongst the 14 inflammatory markers, only hsCRP and hsCRP-to-albumin ratio, Neutrophil-to-Lymphocyte ratio, neutrophil percent-to-albumin ratio, platelet-to-lymphocyte ratio and the SII were not associated with any of SBP, DBP, PP, AIx or PWV. Heart rate was associated with nine of the fourteen inflammatory indices. Monocytes-to-HDL-C, Neutrophil-to-Lymphocytes, Neutrophil percent-to-albumin, Platelet-to-Lymphocytes, and PNI were not associated with HR.

### 3.4 Inflammatory marker X Sex interactions

Associations between the inflammatory markers and BP, and stiffness, were consistently stronger for males compared to females (Table S3 and S4). The significant interactions included those for SBP (sex X mean platelet volume and Neutrophils), for DBP (sex X Neutrophil-to-HDL-C, Neutrophils, and Neutrophil percent-to-albumin), for PP (sex X PNI), for AIx (sex X Neutrophil-to-HDL-C, Monocyte-to-HDL-C, hsCRP, and hsCRP-to-albumin), and for PWV (sex X Lymphocyte count and Neutrophil count).

## 4. Discussion

Within a young and healthy population, our study determined significant associations between novel markers of inflammation derived from routinely measured hematological cell types, including lymphocyte count, monocyte-to-HDL-C ratio and the neutrophil-to-HDL-C ratio, and BP and measures of arterial stiffness (AIx and PP). Our study also provides indicators for the potential mechanisms behind these associations. First, almost all measures of inflammation were significantly associated with heart rate, suggesting increased sympathetic tone may contribute. Second, the positive associations between inflammatory markers and measures of arterial stiffness confirms the latter as a potential mediator of increased BP in young adults (33, 34). Third, the stronger associations observed in males compared to females may either reflect the higher BP and BP variability in males, or true differences arising as the consequence of the many different biological functions impacted by sex including immune cell function (35) and inflammatory disease (36).

Although previous studies have observed associations between inflammation, BP, and arterial stiffness (1, 3, 4), this study provides further evidence of these associations in a young and healthy population and includes novel inflammatory markers. In contrast, CRP was not associated with any measures of either BP or stiffness despite the latter being a predictor of future hypertension in a healthy middle-aged population (2). If the observed BP-inflammatory marker associations reported here reflect the true presence of underlying low-level inflammation and subsequent vascular damage, then WBC-derived markers are potentially more specific than CRP as markers of inflammation related to hypertension. The noticeable increase in BP between age 17 and 22, especially amongst males, supports the notion that the observed associations might be reflecting underlying inflammation and vascular damage related to hypertension within a younger population displaying subtle and gradually increasing systemic low-level inflammation (8).

Our study showed no relationship between NLR, SII or PLR with BP or HR at 17 or 22 years of age. The SII is calculated as the product of the platelet count and the NLR, and has been proposed as an inflammatory marker to evaluate the prognosis of hepatocellular carcinoma (37) and of patients with CVD (38). The NLR has also been used as a separate marker of systemic inflammation and stress in the critically ill (39, 40) and the PLR as a marker in patients with solid tumor (40). The lack of associations in this study may reflect the different populations involved in the original studies, with our healthy and mostly normotensive population differing in both underlying levels of inflammation and disease.

Our findings of an association between lymphocytes, monocytes/HDL-C and neutrophils/HDL-C with arterial stiffness (AIx and PP) support previous literature demonstrating an association between WBCs, vascular function, and arterial stiffness (41, 42). Inflammation has been shown to associate with arterial stiffness and impair vascular function in older adults (1) and increased sympathetic activity may increase and activate T-lymphocytes (1, 19). Our observed associations between inflammatory markers, BP, arterial stiffness, and HR supports both increased sympathetic activity and arterial stiffening as mechanisms for causing increased BP in a young, healthy population.

Our study has several strengths. We measured and adjusted for a wide range of potential confounders including important aspects of diet that can influence BP (sodium, fibre, diet quality), physical activity, WHR, alcohol intake, smoking status, and family history of hypertension. We also used multiple imputation to ensure the inclusion of all subjects in the adjusted models. We measured BP and inflammatory markers at two timepoints which reduced the potential for residual error and considered a wide range of potential markers of inflammation, enabling a comprehensive assessment the best candidates for a simple surrogate measure of inflammation related to hypertension. Finally, our outcomes of BP, HR, AIx, and PWV were aggregated from multiple recordings at each visit, which should also reduce the potential for measurement error and residual confounding.

Our study had several limitations. Although we adjusted for many potential confounders, some of the dietary information used in the adjustment process was obtained at age 20 rather than at ages 17 or 22 when the BP and inflammatory data were obtained. However, since diets of the Raine Study Gen2 adolescents have tracked closely into adulthood (29), adjusting for these measures using age 20 data should still reduce rather than increase the potential for bias. Additionally, we cannot assign causality to our findings due to the observational nature of our study. It is possible that the inflammatory measures are simply corelates for another factor that influences BP, HR, and arterial stiffness, such as physical activity or diet. However, whilst sex, WHR, Western dietary score and METs were consistently associated with many of the inflammatory markers, the observed inflammatory marker-BP associations remained significant after adjusting for sex, WHR, diet quality, physical activity category and metabolic equivalents/day. There remains the potential for residual confounding due to unmeasured confounders such as socioeconomic status, but their influence might be expected to be less than that of sex, diet, physical activity, and adiposity.

Our results demonstrate that lymphocyte count, monocyte-to-HDL-C ratio, neutrophil-to-HDL-C ratio, and platelet count but not hsCRP, SII or NLR are positively associated with BP, arterial stiffness, and HR in young adults. The stronger associations in males compared to females may either reflect the higher BP and variability in the males or the greater potential for inflammatory associated disease amongst males. Novel WBC count and WBC-to-HDL-C derived measures may outperform established measures of inflammation in predicting heightened cardiovascular risk in young adults.

## Acknowledgements

We gratefully acknowledge the Raine Study participants and their families for their continued participation in the study, as well as the Raine Study team for study co-ordination and data collection.

## Institutional Review Board Statement

UWA HREC approval was received on 29 April 2020 and provides a single consolidated approval (RA/4/20/5722) for use of research data and/or biosamples held in the Raine Study data collection. This specific project (Raine Study reference number CAR0713) was reviewed by the Raine Study Scientific Review Committee and approved on 22 of March of 2020. The Southern Adelaide Human Research Ethics Committee approved the project on 29th August 2022 (project ID 4657).

## Informed Consent Statement

Written informed consent from the Raine study Gen2 participants was obtained for each follow-up.

## Data Availability Statement

Data access is subject to restrictions imposed to protect participant privacy. All researchers using Raine Study data must sign a data access agreement stipulating that data may not be released to anyone other than the investigators of the approved project. Additional details regarding data access are available from https://rainestudy.org.au/.

